# Hemispheric Asymmetry Features and Interpretable Machine Learning for Focal Cortical Dysplasia Classification in Drug-Resistant Epilepsy

**DOI:** 10.64898/2026.07.02.26357180

**Authors:** Hoang Nam Dang, Aleena Iraqui

## Abstract

Focal cortical dysplasia (FCD) is a principal cause of pharmacoresistant focal epilepsy, yet its structural MRI signature, subtle cortical thickening, blurring of the gray-white matter junction, is frequently undetected even by experienced neuroradiologists, delaying or precluding referral for curative surgical resection. Here we develop a machine learning pipeline for FCD detection that prioritizes mechanistic interpretability over model complexity. In a subsample of 50 subjects (25 FCD, 25 age-matched controls) drawn from a public structural MRI cohort, we register all scans to a common stereotactic template and derive hemispheric asymmetry features across 48 cortical regions, exploiting the characteristic unilaterality of FCD pathology. Among four classifiers evaluated under leave-one-out cross-validation, an L1-regularized logistic regression achieves the highest accuracy (78%, permutation p=0.02), substantially outperforming tree-based ensembles, which perform at or below chance in this feature-to-sample regime. The fitted model selects a sparse subset of 21 of 96 features, with the largest-magnitude contributions localized to inferior and middle frontal gyri and temporal pole and superior temporal gyrus, regions consistent with the known anatomical distribution of FCD. These findings indicate that hemispheric asymmetry, combined with a sufficiently regularized, interpretable classifier, captures a modest but statistically robust and anatomically grounded signal for FCD detection, offering a transparent complement to existing deep learning approaches for presurgical evaluation.

## 1 Introduction

Epilepsy affects roughly 50 million people worldwide [1], and approximately one-third of cases are resistant to anti-seizure medication. For these patients, epilepsy surgery, which requires resection of the seizure-generating brain tissue, offers the best chance of seizure freedom, but its success depends critically on accurately localizing the epileptogenic lesion prior to surgery [2].

Focal cortical dysplasia (FCD) is among the most common causes of drug-resistant focal epilepsy [3], particularly in children, yet FCD lesions are frequently subtle on structural MRI. They can present as barely perceptible cortical thickening, blurring of the gray-white matter boundary, or minor signal changes that are easily missed on routine visual inspection.

The clinical burden of FCD is disproportionate relative to its prevalence. Marinowic et al. [4] and systematic reviews of drug-resistant epilepsy epidemiology estimate that FCD accounts for over 30% of cases of surgically remediable drug-resistant epilepsy in children [5]. Pharmacoresistance is not merely common in FCD; it is early and often severe: Cohen et al. [6] found that failure of a single antiseizure medication carries substantial risk for subsequent pharmacoresistance, challenging the convention of awaiting failure of two or more agents before referral for surgical evaluation.

The structural MRI signature of FCD has been well characterized in the neuro-radiology literature. Recognized features include focal cortical thickening or thinning, blurring of the gray–white matter junction, gyral and sulcal anomalies, abnormal FLAIR or T2-weighted signal (including the transmantle sign in FCD type IIb), and abnormal interhemispheric asymmetry in structural patterns [7–9]. The last of these, hemispheric asymmetry, occupies a distinctive position among these features: it is directly actionable by radiologists during visual review, and it is grounded in the pathophysiology of FCD, which is almost invariably a focal, unilateral lesion [10]. Conventional MRI protocols at 1.5T or 3T can identify the majority of type II lesions, but sensitivities drop markedly for type I and MRI-negative cases; advanced quantitative and computational post-processing substantially improves detection in these harder subgroups [11].

This detection gap is not hypothetical. In the dataset used for this study, published inter-rater evaluations show that even experienced neuroradiologists correctly identify FCD lesions in only 68% of confirmed cases, non-expert readers in 45%, and laypersons in 27% [12]. Given that a correctly localized lesion is often a prerequisite for surgical candidacy, this gap in visual detection likely contributes to under-referral or delayed referral for surgery in patients who could benefit from it.

Several automated computational approaches have been proposed to close this detection gap. Morphometric Analysis Program (MAP) and voxel-based morphometry toolboxes achieve sensitivities of roughly 64–74% with specificities above 90% on structured research cohorts, but generalization to routine clinical MRI has proved difficult [13]. Deep learning approaches, including three-dimensional convolutional neural networks (CNNs) and, more recently, graph neural networks (GNNs), have pushed reported sensitivities considerably higher: a 2025 JAMA Neurology study of the MELD Graph classifier, trained on data from 23 international epilepsy centers, achieved sensitivity of 81.6% in histopathologically confirmed, seizure-free patients and a positive predictive value of 67%, substantially exceeding a prior surface-based baseline [14]. A 2025 systematic review confirmed that machine-learning classifiers across the literature yield sensitivities of 74–93%, but with wide-ranging specificities (34–100%), and flagged that reported performances may be overly optimistic because most benchmarks compare FCD patients against healthy controls rather than against a realistic clinical mix of patients with other brain abnormalities [11, 15].

A parallel thread of work, closely related to the present study, has formalized hemispheric asymmetry as a computable feature. The original MELD pipeline explicitly constructs interhemispheric asymmetry maps by subtracting right-hemisphere from left-hemisphere vertex values and vice versa, thereby encoding a heuristic that radiologists already apply during visual inspection [10]. Earlier pediatric work by [16] similarly incorporated per-vertex interhemispheric asymmetry alongside cortical thickness and gray–white intensity measures, finding that asymmetry features contributed meaningfully to classifier sensitivity. More recently, Liu et al. [17] extended this logic to radiomic and morphological asymmetry features in pediatric frontal lobe epilepsy, further implicating the contralateral hemisphere as a source of informative contrast signal. These findings collectively motivate the central premise of the current study: that a compact, interpretable feature set built entirely around regional hemispheric asymmetry may retain meaningful discriminative power even when complex model architectures are deliberately avoided.

Machine learning approaches have recently been proposed to address this diagnostic gap. Most notably, the MELD (Multi-centre Epilepsy Lesion Detection) project applies surface-based cortical morphometric features to graph-based and convolutional neural network (CNN) classifiers, enabling automated flagging of putative FCD lesions across large, multi-site cohorts [10]. Such approaches have demonstrated meaningful gains in lesion detection sensitivity relative to unaided visual inspection. However, their clinical translation remains constrained by limited interpretability: a classifier may designate a cortical region as abnormal without providing an anatomically or physiologically grounded rationale for that designation. This opacity is particularly consequential in the context of surgical planning, where neurosurgical teams must critically evaluate, and ultimately assume responsibility for, the biological plausibility of a model’s output before incorporating it into resection planning.

This study takes a narrower, more transparent approach. Rather than pursue state-of-the-art sensitivity with a complex architecture, we build features around hemispheric asymmetry, a heuristic radiologists already use, since FCD lesions are typically unilateral. We pair these features with classical machine learning classifiers whose decisions can be inspected directly: which regions mattered, and which features drove a given prediction. The aim is not to outperform MELD on raw accuracy, but to test whether a simpler, fully interpretable model can still do useful clinical work, and to surface which structural features carry meaningful signal for FCD detection.

## 2 Methods

### 2.1 Dataset

We used the Bonn FCD dataset (OpenNeuro accession ds004199), a publicly available collection of structural MRI scans acquired from 85 patients with histopathologically or radiologically confirmed focal cortical dysplasia and 85 age-matched healthy controls [12]. Scans were acquired under one of two protocols depending on acquisition era: an earlier 1.0 mm isotropic sequence or a subsequent 0.8 mm isotropic sequence. T1-weighted and FLAIR volumes were available for each subject.

### 2.2 Subject selection

From the full cohort, we selected 50 subjects (25 FCD, 25 healthy control) for this analysis, sampled to preserve the age distribution of the full dataset within each diagnostic group. Age at scan did not differ substantially between groups in the selected subsample (FCD mean age 6.16 years, HC mean age 7.12 years), consistent with the age distribution of the full 170-subject cohort (FCD mean 6.19 years, HC mean 7.07 years).

### 2.3 Preprocessing

T1-weighted volumes were skull-stripped using a deep learning-based brain extraction model implemented in ANTsPyNet [18], then registered to the MNI152NLin2009cAsym template (1 mm isotropic) [19] via symmetric diffeomorphic normalization (SyN) as implemented in ANTsPy. Registration quality was visually inspected in a representative subset of subjects across both acquisition protocols, with attention to cortical boundary alignment and ventricular correspondence between the normalized volumes and the template. Representative results of skull-stripping are shown in Fig. 1, and registration accuracy for the same subject is shown in Fig. 2.

**Fig. 1.**
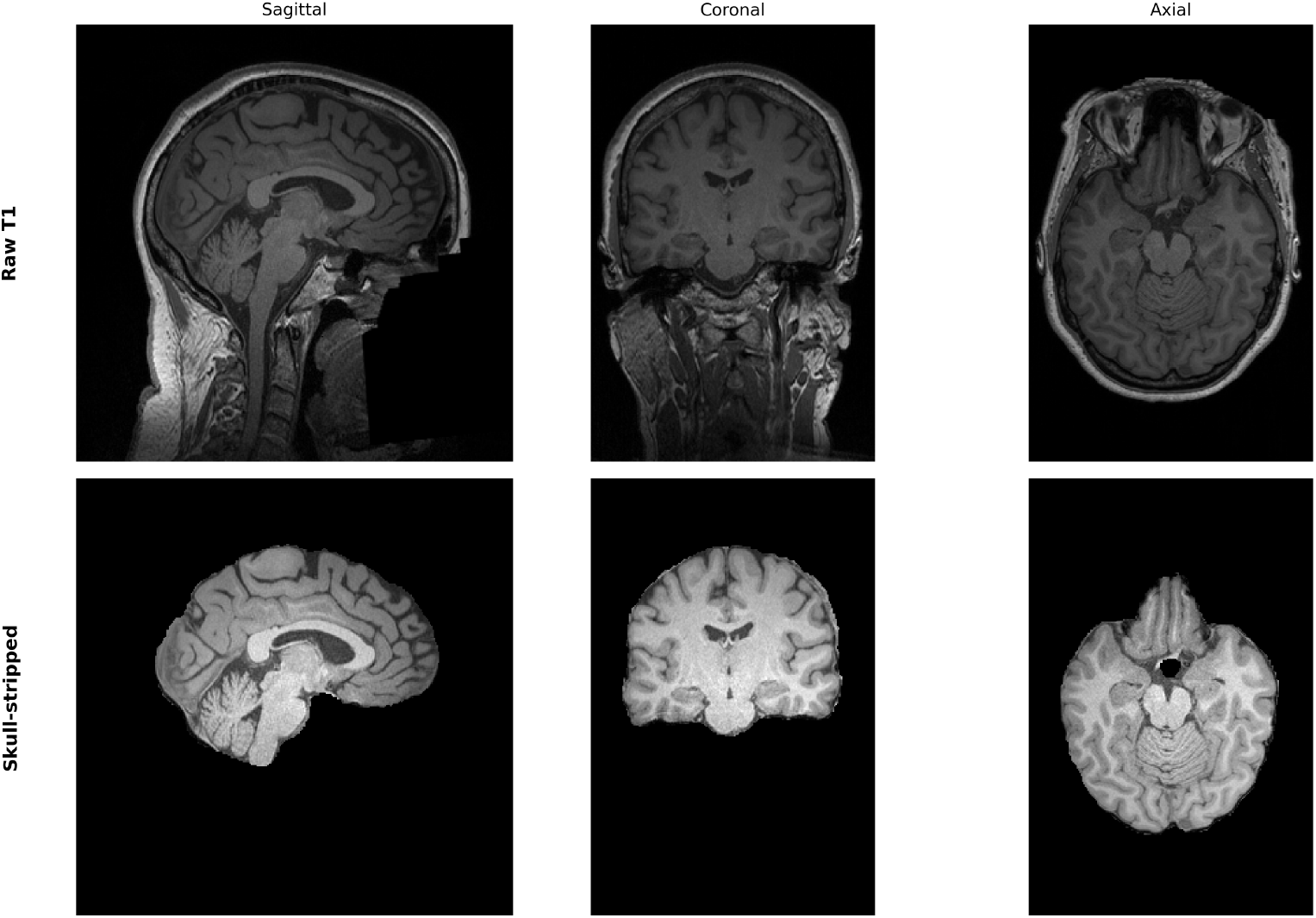
Representative preprocessing for a single subject (sub-00002). Top row: raw T1-weighted volume. Bottom row: skull-stripped volume following deep learning-based brain extraction, shown in sagittal, coronal, and axial planes.

**Fig. 2.**
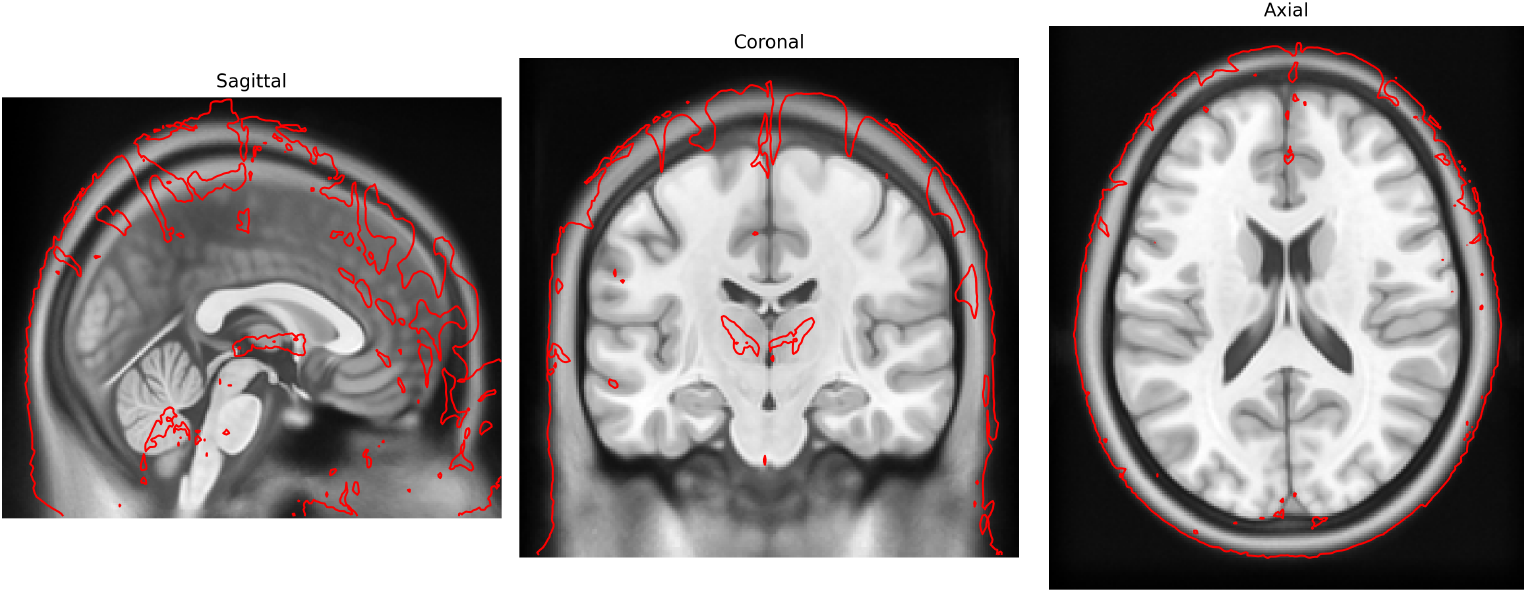
Registration quality control for the same subject. Red contours indicate the boundary of the registered, skull-stripped brain overlaid on the MNI152NLin2009cAsym template (grayscale), shown in sagittal, coronal, and axial planes.

### 2.4 Regional parcellation

Normalized brains were parcellated using the Harvard-Oxford cortical atlas, which defines 48 cortical regions [20]. Because the atlas is distributed in a different standard-space variant than our registration target, it was first resampled onto the target grid using nearest-neighbor interpolation, which preserves discrete label values and avoids the spurious intermediate labels that continuous interpolation would introduce. Each region, defined bilaterally in the atlas, was then divided into separate left and right components according to voxel position relative to the anatomical midline. The midline was determined from the image affine and orientation metadata rather than assumed to coincide with the volumetric center, and the resulting hemisphere assignments were verified against the orientation codes, as illustrated in Fig. 3 for a representative region. This procedure yielded 96 unilateral regions, organized as 48 homologous left-right pairs.

**Fig. 3.**
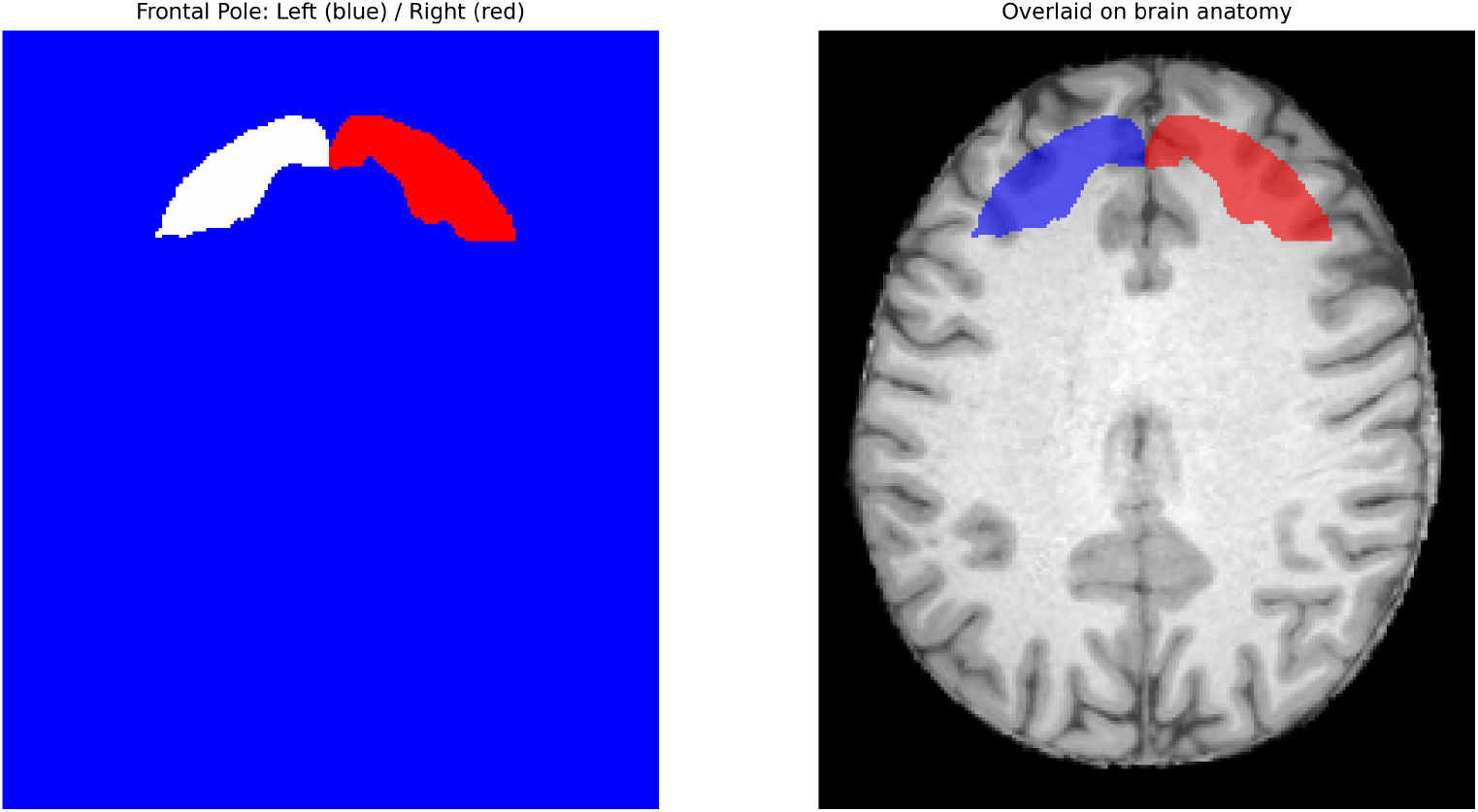
Verification of hemisphere assignment following atlas splitting, shown for the frontal pole region. Left (blue) and right (red) components after splitting the bilateral Harvard-Oxford label along the anatomical midline, shown in isolation (left panel) and overlaid on subject anatomy (right panel).

### 2.5 Feature extraction

For each of the 48 homologous region pairs, we quantified interhemispheric asymmetry, motivated by the observation that focal cortical dysplasia is typically a unilateral pathology and therefore expected to manifest as a departure from the near-symmetry of the healthy brain. We computed two complementary asymmetry measures. The first captured differences in mean signal intensity between homologous regions, and the second captured differences in intensity dispersion, quantified as the standard deviation of voxel intensities within each region. Each measure was expressed as a normalized asymmetry index of the form 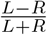, where *L* and *R* denote the corresponding statistic computed over the left and right members of a region pair, respectively. This normalization bounds each feature to the interval [*−*1, 1] and renders it invariant to global scaling of intensity, mitigating between-subject and between-protocol differences in absolute signal. Combining two measures across 48 region pairs produced a 96-dimensional feature vector per subject.

### 2.6 Classification

We framed FCD detection as a binary classification problem, distinguishing patients from healthy controls on the basis of the 96 asymmetry features. Rather than commit to a single learning algorithm, we evaluated four classifiers spanning distinct inductive biases: a random forest (200 trees) [21], gradient boosted decision trees [22], an L1-regularized logistic regression [23], and a support vector machine with a linear kernel [24]. This comparison was motivated by the high ratio of features to observations in our data, a regime in which model complexity and the propensity to overfit differ substantially across algorithms, and in which no single method can be assumed optimal a priori. Features supplied to the logistic regression and support vector machine were standardized to zero mean and unit variance prior to fitting, as both are sensitive to feature scaling; the tree-based ensembles, which are invariant to monotonic transformations of individual features, were applied to the unscaled features.

Given the modest sample size, we assessed generalization performance using leave-one-out cross-validation, in which the model is trained on all but one subject and evaluated on the held-out individual, with the procedure repeated so that each subject serves once as the test case. This scheme maximizes the data available for training at each iteration and yields a nearly unbiased estimate of expected performance, an important consideration when a conventional held-out test partition would comprise only a handful of subjects and produce an unstably estimated accuracy.

### 2.7 Statistical validation

Classification accuracy, considered in isolation, is difficult to interpret when the number of features approaches or exceeds the number of observations, since flexible models can achieve apparently high accuracy by fitting sample-specific structure that does not generalize. To establish whether the performance of our best classifier exceeded that attributable to chance, we conducted a permutation test [25]. The diagnostic labels were randomly permuted across subjects, severing any genuine association between the asymmetry features and group membership while preserving the marginal class distribution, and the complete cross-validation procedure was repeated on the permuted labels. Iterating this process 100 times generated an empirical null distribution of cross-validated accuracies. We report the permutation-based *p*-value as the proportion of permutations whose accuracy equaled or exceeded the accuracy obtained under the true labeling.

## 3 Results

### 3.1 Sample characteristics

The analyzed cohort comprised 50 subjects, evenly divided between FCD patients (*n* = 25) and healthy controls (*n* = 25). The two groups were closely matched in age at scan (FCD, mean 6.16 years; control, mean 7.12 years), and this distribution mirrored that of the full 170-subject dataset from which the subsample was drawn (FCD, mean 6.19 years; control, mean 7.07 years), indicating that the subsampling procedure preserved the age structure of the source population and did not introduce a systematic age difference between groups.

### 3.2 Classification performance

We evaluated four classifiers on the 96-dimensional asymmetry feature set under leave-one-out cross-validation (Table 1, Fig. 4). Performance varied substantially across algorithms. The L1-regularized logistic regression achieved the highest accuracy at 78.0% (39 of 50 subjects correctly classified), followed by the linear support vector machine at 68.0% and the random forest at 66.0%. The gradient boosted ensemble performed at 40.0%, below the 50% expected under chance for this balanced problem.

**Table 1.** Cross-validated classification accuracy for the four evaluated classifiers under leave-one-out cross-validation.

| Classifier | Accuracy (%) | Correct / Total |
| --- | --- | --- |
| L1-regularized logistic regression | 78.0 | 39 / 50 |
| Linear support vector machine | 68.0 | 34 / 50 |
| Random forest | 66.0 | 33 / 50 |
| Gradient boosted trees | 40.0 | 20 / 50 |

**Fig. 4.**
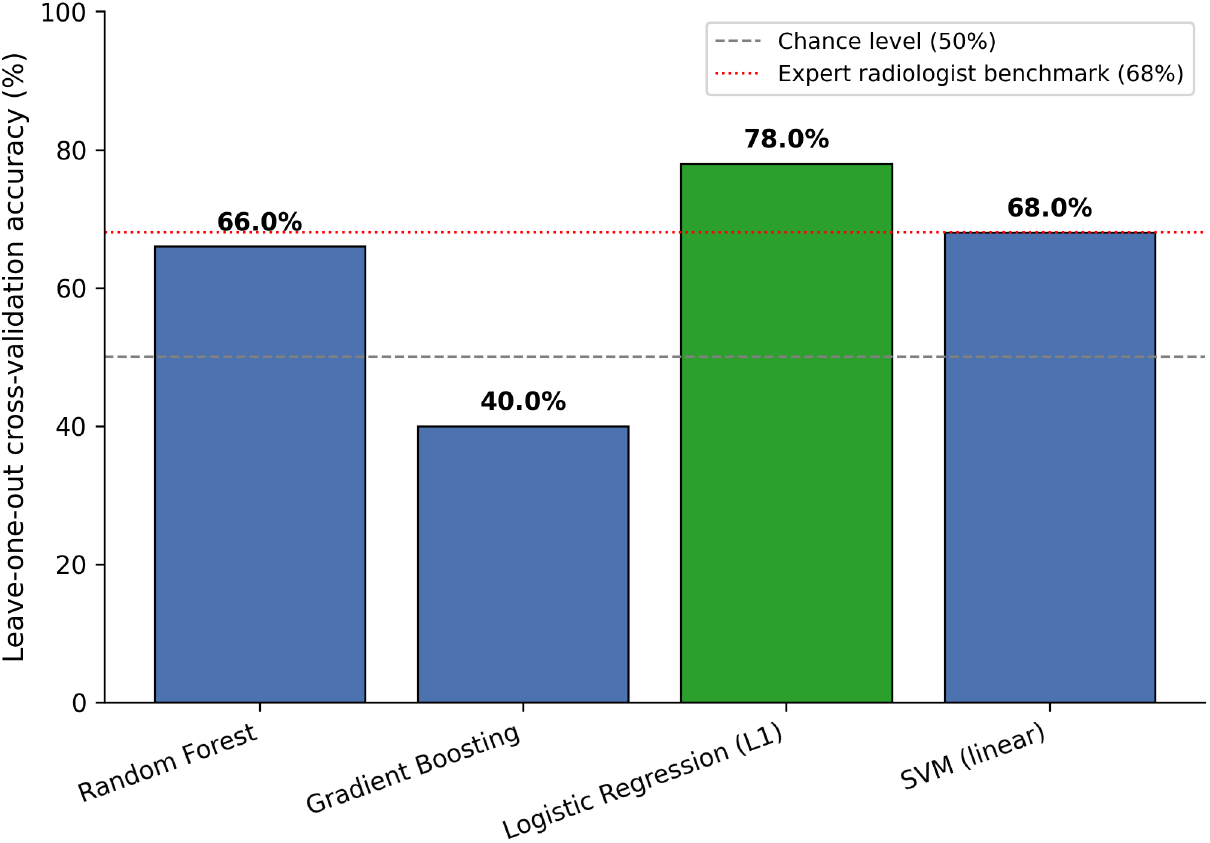
Cross-validated classification accuracy across four classifiers. Dashed gray line indicates chance level (50%); dotted red line indicates the reported sensitivity of expert neuroradiologists on this dataset (68%).

This ordering is consistent with the high ratio of features to observations in the data. The two models that performed best, logistic regression and the linear support vector machine, impose strong constraints on model complexity, through L1 regularization and margin maximization respectively, whereas the gradient boosted ensemble, the most flexible of the four, performed worst. Rather than an incidental observation, this pattern indicates that in the present regime, restricting model capacity is beneficial and that highly flexible learners overfit sample-specific structure that does not generalize.

### 3.3 Statistical significance

To determine whether the accuracy of the best-performing classifier exceeded chance, we subjected the logistic regression to a label-permutation test with 100 permutations. The resulting null distribution of cross-validated accuracies centered near chance (mean 51.9%, standard deviation 12.2%), confirming that the procedure recovered no systematic signal when the association between features and diagnosis was destroyed. The accuracy obtained under the true labeling (78.0%) exceeded all but two of the permuted accuracies, corresponding to a permutation-based *p*-value of 0.02 (Fig. 5). The maximum accuracy observed across all permutations was 82.0%, indicating that although the true result is statistically distinguishable from chance, the null distribution retains appreciable spread at this sample size.

**Fig. 5.**
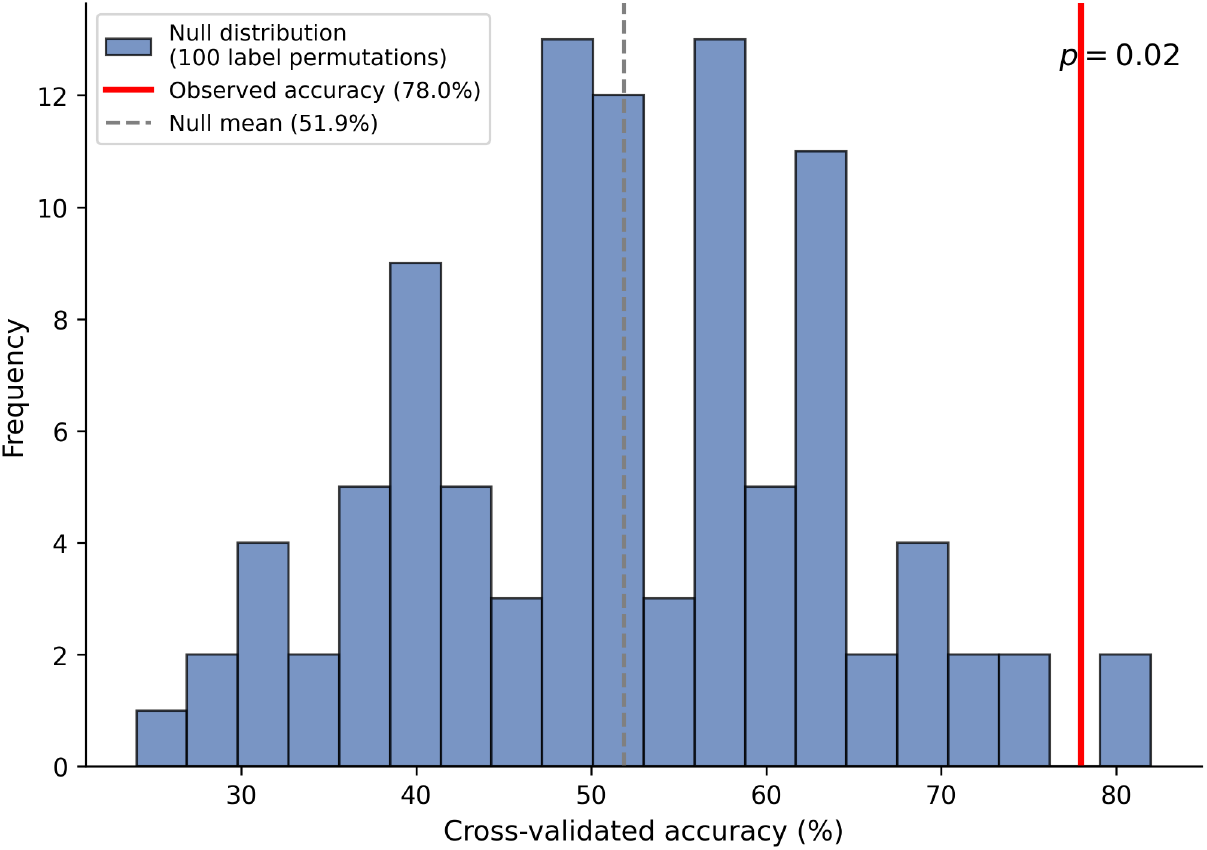
Empirical null distribution of cross-validated accuracy under 100 random permutations of diagnostic labels, using the L1-regularized logistic regression. The observed accuracy under the true labeling (78.0%, red line) exceeds all but two permutations (*p* = 0.02).

### 3.4 Feature interpretation

A principal motivation for the L1-regularized model is that it performs embedded feature selection, retaining only features that contribute to the decision boundary and setting the remaining coefficients to zero. Of the 96 asymmetry features, 21 received non-zero weights in the fitted model (Table 2). The features carrying the largest coefficient magnitudes localized predominantly to the frontal and temporal lobes: the inferior frontal gyrus (pars triangularis) and middle frontal gyrus in the frontal lobe, and the temporal pole and superior and middle temporal gyri in the temporal lobe (Fig. 6). This anatomical distribution is concordant with the established epidemiology of FCD, in which frontal and temporal locations predominate, and provides converging evidence that the classifier’s discriminative signal reflects genuine pathology-related structure rather than arbitrary sample-specific variation.

**Table 2.**
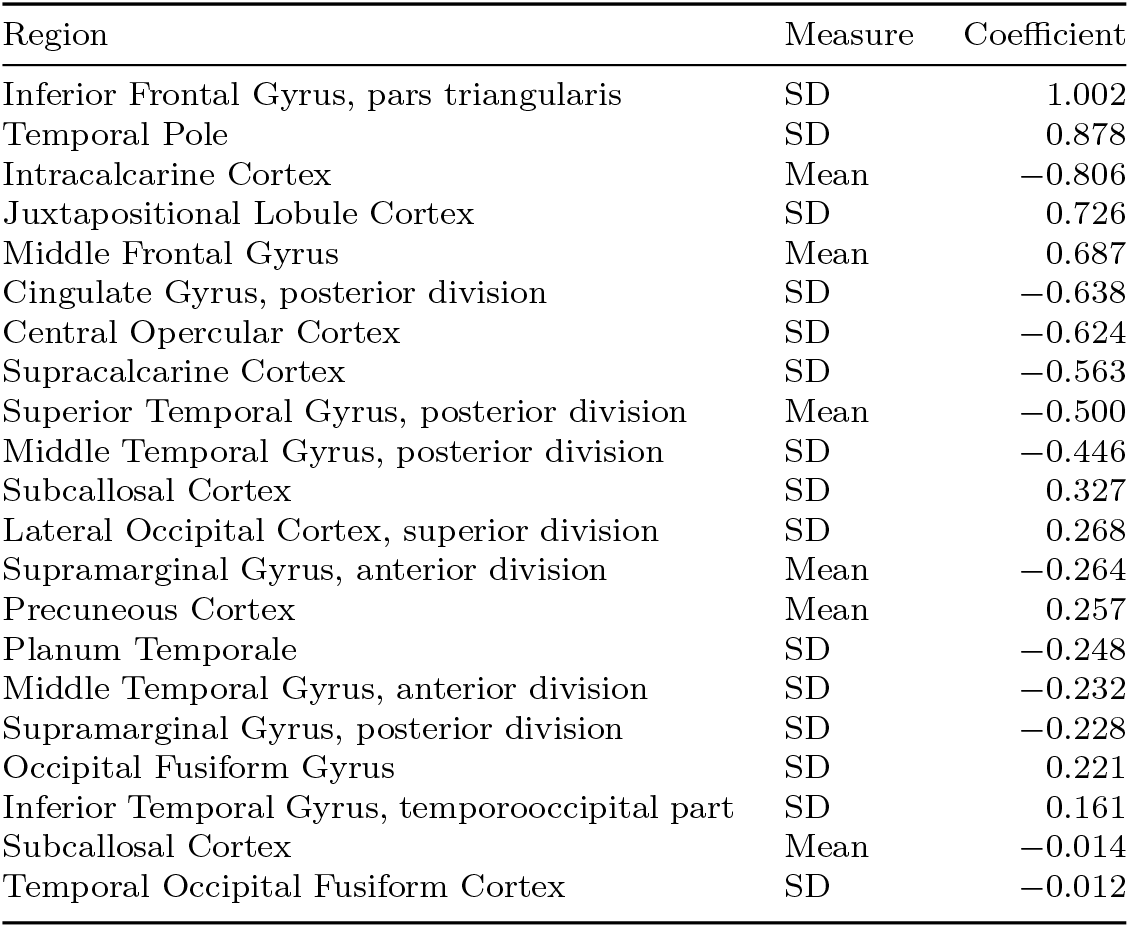
Features retained by the L1-regularized logistic regression, ordered by absolute coefficient magnitude. Positive coefficients indicate association with the FCD class, negative with the control class. Asymmetry measures are the normalized left-right index of regional mean intensity (Mean) or intensity standard deviation (SD).

**Fig. 6.**
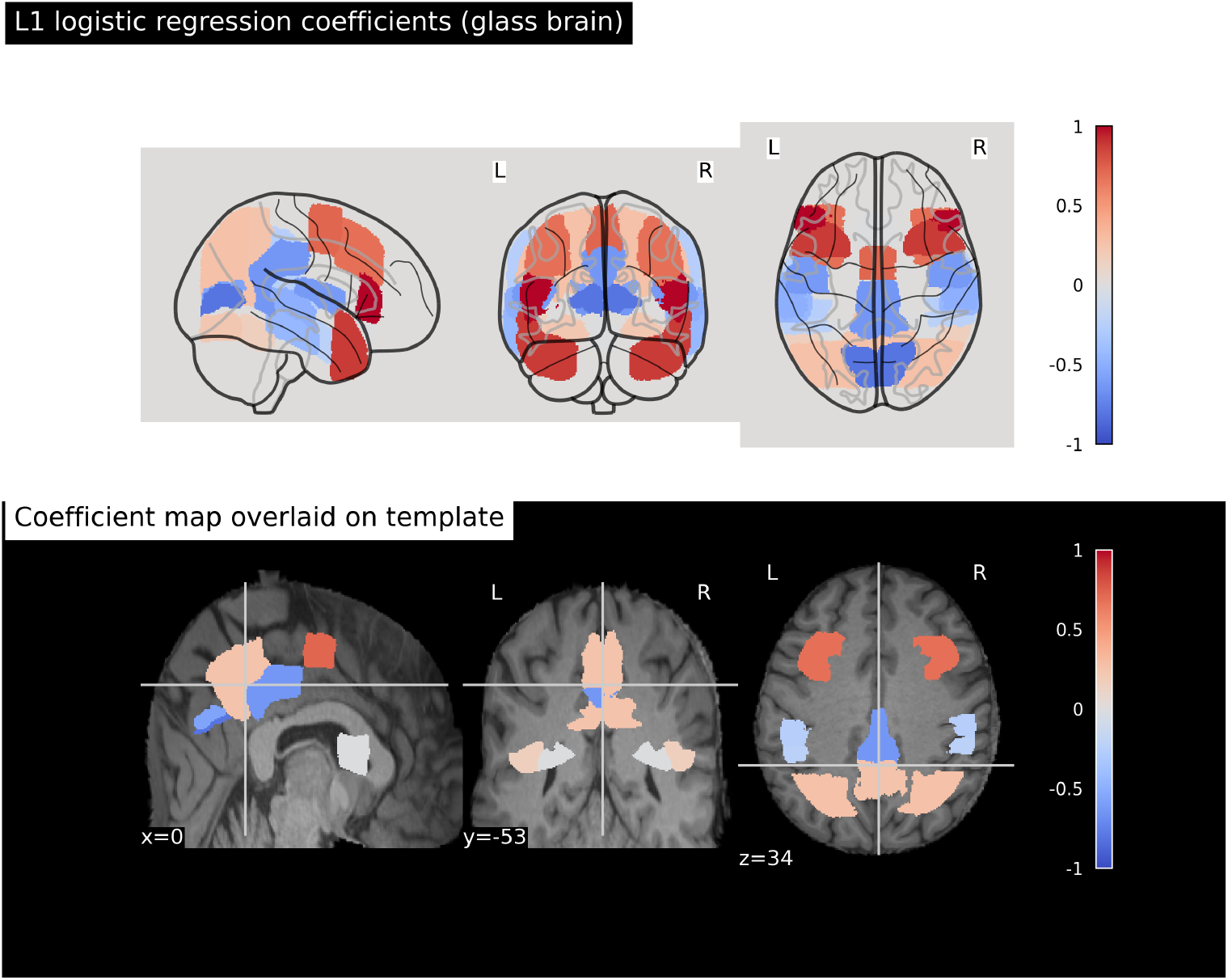
Spatial distribution of L1 logistic regression coefficients. Glass brain (top) and orthogonal slice views overlaid on the template (bottom) show regions retained by the sparse model, colored by coefficient sign and magnitude. Warm colors indicate positive association with the FCD class; cool colors indicate negative association.

Several selected features localized to occipital and medial regions, including the intracalcarine and supracalcarine cortices and the subcallosal cortex, whose relationship to FCD is less clearly established. We interpret these selections with caution. At the present feature-to-sample ratio, some degree of sample-specific selection is expected even under regularization, and we do not claim that every retained feature reflects generalizable pathology. Both the intensity-mean and intensity-dispersion measures contributed to the selected set, suggesting that regional differences in signal heterogeneity, and not only in mean signal, carry discriminative information.

## 4 Discussion

Across four classifiers, an L1-regularized logistic regression distinguished FCD patients from healthy controls with 78% accuracy under leave-one-out cross-validation, a result confirmed by permutation testing to exceed chance (*p* = 0.02). Though modest, this accuracy surpasses the reported sensitivity of expert neuroradiologists on this dataset [12], and it is obtained from a transparent model whose decision rule can be inspected directly.

The comparison across classifiers is itself informative. The two best-performing models constrained their own complexity, through L1 regularization and margin maximization, whereas the most flexible model, gradient boosted trees, performed below chance. Where features nearly double the number of observations, this is the expected signature of overfitting, and it suggests that model parsimony here serves generalization, not only interpretability.

The features retained by the sparse model offer partial construct validity. The largest-magnitude coefficients localized to frontal and temporal regions, the predominant anatomical sites of FCD [26, 27], suggesting the discriminative signal is at least partly grounded in disease-relevant anatomy. Several retained features, however, localized to occipital and medial regions with no clear relationship to FCD; we offer no post hoc rationale for these and regard them as provisional, consistent with the sample-specific selection expected at this scale. Our approach also differs in philosophy from established pipelines such as MELD, which apply surface-based morphometry and deep learning to achieve high sensitivity across large multi-site cohorts [10, 14]. We do not position our method as a competitor on sensitivity, but as evidence that a substantial fraction of the FCD signal is accessible to a simple, fully transparent model useful both as an interpretable adjunct and as a baseline for quantifying the added value of more complex methods.

Several limitations bound these results, and several plausible confounders remain uncontrolled. Most fundamentally, the analysis rests on 50 subjects from a single dataset; the reported accuracy and the specific features selected require validation in larger and independent cohorts before any clinical inference is warranted. We did not perform nested cross-validation or an external hold-out, so the reported accuracy may still be optimistic. Although we age-matched the groups, we did not control for sex, handedness, scanner or acquisition protocol, or total brain volume, any of which could contribute residual asymmetry signal unrelated to pathology; the two acquisition protocols in particular could induce systematic intensity differences that our normalization only partially removes. Our reliance on registration to a symmetric template also means residual misregistration could masquerade as anatomical asymmetry, and we assessed registration quality visually rather than quantitatively. During development we identified and corrected two further pitfalls that we report because they illustrate failure modes easy to commit and consequential: a volumetric asymmetry feature that was invariant across subjects, because region volumes computed from a shared template atlas necessarily coincide once all brains are warped to it, and a spurious age imbalance in an early exploratory subsample that inflated apparent performance until corrected by age-matched sampling [28]. Finally, we used only T1-weighted images despite the availability of FLAIR, on which FCD lesions are frequently more conspicuous [9, 29], and a single cortical atlas at a single spatial scale; both constrain sensitivity and are natural targets for future work.

## 5 Conclusion

We have presented an interpretable machine learning pipeline for FCD detection that derives interhemispheric asymmetry features from template-normalized structural MRI and classifies them with deliberately constrained models. On a balanced, age-matched sample of 50 subjects, a sparse logistic regression distinguished patients from controls with 78% accuracy, a result that was statistically significant by permutation testing and anchored to frontal and temporal features consistent with known FCD anatomy. The work is exploratory in scale and makes no claim to clinical readiness, but it supports a specific and useful proposition: that a transparent, low-complexity model can extract a meaningful and anatomically plausible signal for FCD from structural asymmetry alone.

Several directions follow naturally. The most immediate is to incorporate FLAIR-derived features and to scale the analysis to the full dataset and, ultimately, to independent multi-site cohorts, both of which would test the stability of the present findings and the specific features implicated. Beyond confirmation, a natural extension is to move from subject-level classification toward lesion localization, mapping which regional asymmetries drive an individual prediction, so that an interpretable model of this kind could serve not only as a screening aid but as a means of directing radiological attention within a given scan. We view the present study as a baseline and a starting point for that trajectory rather than an endpoint.

## Data Availability

All data produced are available online at https://openneuro.org/datasets/ds004199/versions/1.0.6

## Notes

### Competing Interest Statement

The authors have declared no competing interest.

### Author Declarations

The study will only use openly available human data that were originally located at https://openneuro.org/datasets/ds004199/versions/1.0.6

